# Real-time PCR assays for detection and quantification of early *P. falciparum* gametocyte stages

**DOI:** 10.1101/2021.03.28.21254192

**Authors:** Amal A.H. Gadalla, Giulia Siciliano, Ryan Farid, Pietro Alano, Lisa Ranford-Cartwright, James S McCarthy, Joanne Thompson, Hamza Babiker

## Abstract

**Introduction:** The use of reverse transcription, quantitative qRT-PCR assays for detection and quantification of late gametocyte stages has revealed the high transmission capacity of the human malaria parasite, *Plasmodium falciparum*. A full understanding how the parasite adjusts its transmission in response to varying in-host environmental conditions during natural infections requires simultaneous quantification of early and late gametocytes. Here, we describe qRT-PCR assays that are specific for detection and quantification of early-stage gametocytes of *P. falciparum*.

**Methods:** The assays are based on expression of known early gametocyte genes (*pfpeg4, pfg27, pfge1, pfge3* and *pfgexp5*). The specificity of the qRT-PCR assays was tested using purified stage II and stage V gametocytes. These validated assays were used with qRT-PCR assays targeting late stage (*pfs25)* and all-stage (*pfs16*) gametocyte-specific transcripts to quantify gametocytes in natural *P. falciparum* infections and in a controlled human clinical infection study.

**Results:** The relative expression of *pfpeg4, pfg27* and *pfge3*, but not of *pfge1* and *pfgexp5*, was significantly higher in purified stage II compared to stage V gametocytes, indicating early gametocyte specificity. In natural infections, 71.2% of individuals had both early and late gametocyte transcripts (*pfpeg4*/*pfg27* plus *pfs25*), 12.6% harboured only early gametocytes transcripts (*pfpeg4*/*pfg27)*, and 15.2% had only late gametocytes transcripts (*pfs25*). In natural infections, the limit of detection was equivalent to 190 and 390 gametocytes/mL blood for *pfpeg4* and *pfg27*, respectively. In infected volunteers, transcripts of *pfpeg4* and *pfg27* were detected shortly after the onset of blood stage infection, demonstrating the specificity of the assays.

**Conclusion:** The *pfpeg4* and *pfg27* qRT-PCR assays can be used specifically to quantify circulating immature gametocytes. Quantification of early gametocytes will improve understanding of epidemiological processes that modulate *P. falciparum* transmission and enhance the evaluation of transmission blocking interventions.

## 1. Introduction

The malaria parasite *Plasmodium falciparum* continues to be a major cause of human disease despite mounting control measures. Central to this success is its ability to undergo sexual development and efficient transmission between human hosts through its mosquito vector. In *P. falciparum*, sexual development occurs when asexual blood stage parasites exit the proliferative cycle and develop into schizonts containing merozoites committed to sexual differentiation (Bruce et al., 1990). These merozoites invade erythrocytes and differentiate into the sexual forms, gametocytes, over a period of approximately 10 days, passing through five morphologically distinct stages (I to V) (Carter and Miller, 1979). A second direct path to commitment to gametocytogenesis may also occur shortly after invasion of the erythrocyte by an uncommitted merozoite (Bancells et al., 2019). While very early and late-stage gametocytes (stages I and V) are present in the peripheral circulation, immature gametocytes (stage II-IV) withdraw from peripheral circulation and sequester in tissue microvasculature (Smalley et al., 1981, Joice et al., 2014, Aguilar et al., 2014, Obaldia et al., 2018).

Stage I gametocytes are morphologically indistinguishable from pigmented asexual trophozoites, and thus cannot be identified in peripheral blood by microscopic examination. Transcriptome analyses has led to the identification of transcripts upregulated before morphological differentiation into stage II gametocytes. These include *pfmdv-1 (pfpeg3), pfgdv1, pfge1*-*3* (Silvestrini et al., 2005, Eksi et al., 2005, Eksi et al., 2012) and *pfpeg4* (Joice et al., 2013), which encodes the membrane protein ETRAMP 10.3 (Silvestrini et al., 2005, MacKellar et al., 2010, Buchholz et al., 2011). Further analyses of the transcriptomes of *P. falciparum* isolates have demonstrated that transcripts of *pfge1, pfge2* and *pfg27* cluster away from those of late stage-specific genes *pfs25, pfs28* and *pfs47*, suggesting that they are expressed exclusively in early stage gametocytes (Eksi et al., 2012). In addition, Tiburcio et al., (2015) described *pfgexp5* as the earliest gametocyte-specific gene, expressed 14 hours post-invasion. This is consistent with the detection of *pfgexp5* in blood samples taken at early time points following experimental infection of volunteers with *P. falciparum* (Farid et al., 2017).

The development of RT-PCR assays based on late sexual stage-specific mRNA transcripts has enabled epidemiological surveys and revealed the transmission potential of *P. falciparum* that drives its success in the face of continued control efforts (Babiker et al., 1999, Nassir et al., 2005, Schneider et al., 2006, Menegon et al., 2000). These surveys have identified a range of epidemiological correlates that modulate transmission, including drug treatment, anaemia, mixed species infection and multiplicity of genotypes (Bousema et al., 2004, Babiker and Schneider, 2008, Mawili-Mboumba et al., 2013, Ouédraogo et al., 2009). These epidemiological findings are consistent with the predictions of the evolutionary hypotheses that the interplay between asexual replication and transmission is altered by a changing in-host environment (Reece et al., 2008, Carter et al., 2013). However, to disentangle the effect of these factors on transmission success, there is a need for assays that can detect and quantify very early gametocytes close to the point of commitment.

Here, we describe sensitive qRT-PCR assays for quantification of early stage gametocytes, that complement existing late-stage assays, to enable the analysis of epidemiological factors that drive *P. falciparum* transmission success, and robust assessment of control strategies targeting sexual stages. The assays were validated using *in vitro* purified early and late *P. falciparum* gametocytes and their robustness was tested using samples from natural infections and from experimentally-infected volunteers.

## 2. Methods

### 2.1. Asexual and purified stage II and V gametocyte samples

*P. falciparum* clone 3D7A was cultivated as described by Trager and Jensen (1976) with minor modifications. Briefly, parasites were maintained in human type O-positive RBCs at 5% haematocrit (Hct) in RPMI 1640 medium supplemented with 25 mM HEPES (Sigma), 50 µg/ml hypoxanthine and with the addition of 10% (v/v) naturally-clotted heat-inactivated 0+ human serum (Interstate Blood Bank, Inc.). The cultures were maintained at 37°C in a standard gas mixture consisting of 3% O_2_, 5% CO_2_ and 92% N_2_. To obtain synchronous gametocytes, an asexual culture (0.5-1% starting parasitaemia) was grown without further dilution as described (D’Alessandro et al., 2013). Induced gametocyte cultures were supplemented with 50 mM N-acetylglucosamine (NAG; Sigma-Aldrich) to clear residual asexual parasites and obtain a virtually pure gametocyte culture (Gupta et al., 1985). To obtain the stage II gametocyte sample, two days after NAG treatment gametocytes were inspected on Giemsa-stained smears to assess maturation, and purified by centrifugation through a discontinuous Percoll gradient, as described in Silvestrini et al. (2010). Gametocytes were examined in a counting chamber for quantification and control of contamination. Stage IV and V gametocytes were not detectable. A small fraction of residual unhealthy/dead trophozoites and uninfected RBCs was detectable. To obtain the stage V gametocyte sample, gametocytes were induced as described before (D’Alessandro et al., 2013), and 12 days after NAG treatment gametocytes were purified by MACS Separation Columns CS (Miltenyi Biotec) (Ribaut et al., 2008) and examined as above. Immature sexual stages and asexual stages could not be detected. In both cases the enriched parasite preparations were pelleted and frozen in liquid nitrogen. Samples containing mixed-stage asexual parasites, purified stage II gametocytes, and purified stage V gametocytes were used to examine the stage specificity of the candidate early gametocyte genes and to establish the mathematical relationship between transcript numbers of the candidate genes and gametocyte counts (more details are given in section 2.6).

### 2.2. Natural *P. falciparum* infections

A total of 250 samples were examined from an existing collection, stored at −80°C as packed RBCs. The samples were obtained from patients with uncomplicated *P. falciparum* malaria from Asar village, eastern Sudan, with ethical clearance from the Ministry of Health, Sudan (Ali et al., 2006). The samples were employed in the current study to investigate whether the expression of early gametocyte markers is: 1) detectable in natural infections; 2) detectable specifically in a subset of samples that were *pfs16* positive (early and late gametocyte markers) and *pfs25* negative (late gametocyte marker); 3) associated with total parasitaemia.

### 2.3. Experimentally infected human volunteers

Samples from experimentally-infected human volunteers, who had participated in a previously published clinical trial (Farid et al., 2017, Pasay et al., 2016), were utilized to study the stage-specificity of the gametocyte markers *in vivo*. Ethical approval of the study was obtained from QIMR Berghofer Human Research Ethics Committee. The volunteer cohort has been previously described. In brief, it comprised malaria-naïve, healthy males and non-pregnant females, aged 18-50 years. Infections were induced using approximately 1800 viable *P. falciparum*–infected human erythrocytes of clone 3D7. Volunteers were then treated with 480 mg of piperaquine (which affects only asexual stages) on Day 8 (D08) post-infection, when parasitaemia reached a predetermined threshold of >1,000 parasites/mL. In the current study, samples were examined from two of these volunteers who showed a relatively high post-treatment parasitaemia, and high levels of *pfs25* expression at or beyond D11 post-infection (Farid et al., 2017). Ten time points were selected for each volunteer, between D07 to D09 post-infection. This represents a suitable window for interrogation for early gametocytes, as parasitaemia started to increase at D07, with limited possibility for the presence of late gametocytes in the initial inoculum as discussed (Farid et al., 2017).

### 2.4. Total parasite density quantification

Parasite density was quantified as parasites/mL of blood using absolute quantification of the *18S rRNA* gene by qPCR assay (Hermsen et al., 2001, Gadalla et al., 2016). *18S rRNA* copy numbers were converted to parasite numbers using a calibration curve from *P. falciparum* clone 3D7 parasite DNA with a range of 0.14 to 138938 parasites/μL of DNA (Nwakanma et al., 2009). The *18S rRNA* qPCR amplification efficiency was 97.7% (se 0.01%). Quantification was carried out in duplicate with 0.35 standard deviation between replicates. Quantification of parasite density in the volunteers’ samples has been described elsewhere (Farid et al., 2017).

### 2.5. DNA, RNA extraction, purification and cDNA preparation

For samples from naturally-infected patients, DNA and RNA were extracted from 100 µL and 50 µL of packed RBCs, using the QIAamp DNA mini kit (Qiagen) and SV Total RNA Isolation System (Promega, UK) respectively. For RNA purification, all samples were treated with a unified concentration of 1 unit of RQ1 RNase-Free DNase (Promega, UK) per 8 µL of RNA sample to remove any genomic DNA (gDNA) carryover. Then, purified RNA samples were checked for gDNA by*18S rRNA* qPCR assays. Pure RNA samples were converted to cDNA using the High-Capacity cDNA Reverse Transcription Kit with random primers (Thermo Fisher, UK). For the volunteer cohort, RNA was extracted from 800 µL of whole blood and cDNA was prepared as described (Farid et al., 2017).

### 2.6. qRT-PCR assays for early and late gametocytes

#### 2.6.1. Oligonucleotide design

Full gene names and accession numbers are provided in supplementary Table S1. qRT-PCR primers and TaqMan dual-labelled probes were designed within exon sequences, avoiding polymorphic regions to ensure reproducibility of the experiment for field isolates (Table S2). BLAST alignments (Ensembl.org, 2014, PlasmoDB.org, 2014) against sequences available in PlasmoDB.org (2014) showed that the primers and probe sets had 100% identity with *P. falciparum* target genes, and no high identity alignment with other human *Plasmodium* species or other *P. falciparum* genes. This was further confirmed by the absence of amplification when primers and probes were used in PCR assays using *Plasmodium vivax, Plasmodium malariae* and *Plasmodium ovale* gDNA. Probe design for *pfgexp5* was described previously by Farid et al. (2017).

#### 2.6.2. Optimization

A Taqman assay was designed and optimised for each of the putative early gametocyte-specific genes (*pfpeg4, pfg27, pfge1, pfge3* and *pfgexp5*), for one late gametocyte-specific gene (*pfs25*), for one pan-gametocyte marker (*pfs16*), and for one reference gene (*pf40S*). Primer concentrations for the optimised qRT-PCR reactions were 0.3 µM (*pfpeg4, pfg27, pfge1* and *pfs25*), 0.6 µM (*pfgexp5*), and 1.2 µM (*pfge3*). Probe concentrations used were 0.1 µM in the *pfpeg4, pfge1, pfge3, pfgexp5* and *pfs25* qRT-PCR assays and 0.2 µM in the *pfg27* and *pf40S* assays. The final reaction contained 1X TaqMan Universal PCR Master Mix, No AmpErase UNG, ABI. The temperature profile was; 2 min at 50°C, 10 min at 95°C, and then 45 cycles of 15 sec at 95°C and 1 min at 60°C. Quantification was carried out in duplicate with 0.35 standard deviation between replicates.

#### 2.6.3. Generation of qRT-PCR standard curves

To provide enough material for qRT-PCR optimization, establishment of sensitivity and for sample quantification assays, standard curves were based on cDNA that was transcribed *in vitro* from DNA templates, hereafter named ivcDNA. DNA templates for each gene were prepared by PCR amplification of the target regions from *P. falciparum* clone 3D7 incorporating primers containing SP6 or T7 RNA polymerase promoter sequences (Table S2). Amplicons were purified from excess reagent and primers using Wizard SV Gel and PCR Clean-Up System (Promega) as described by the manufacturer, and the concentration was determined using nanodrop (NanoDrop Spectrophotometer, ND1000, Thermo Fisher Scientific). The number of molecules of DNA amplicon was calculated using the equation (ng DNA × 6.022 × 10^23^) / length (bp) of the DNA template × 10^9^ x 650. Purified DNA amplicon was then transcribed to RNA using Riboprobe Combination System-SP_6_/T_7_ RNA Polymerase (Promega) as described by the manufacturer. gDNA was removed from RNA using 1 unit of DNase treatment (RQ1 DNase, Promega) per 8 µL of RNA and qPCR was carried out to confirm the purity of the RNA prior to cDNA preparation. ivcDNA was prepared by RNA reverse transcription using High-Capacity cDNA Reverse Transcription Kit, Applied Biosystems (ABI), USA as described by the manufacturer. The concentration of the ivcDNA was calculated based on the initial concentration of DNA amplicon and incorporating the dilution factors occurred during the processes above (DNA transcription, RNA purification and RNA reverse transcription). ivcDNA was then used to generate standard curves to assess qRT-PCR efficiency and sensitivity and to quantify the transcript number of the early gametocyte candidate genes in clinical samples.

#### 2.6.4. Quantification of gametocyte gene expression and gametocyte numbers

The limit of quantification (LoQ) for the qRT-PCR assays was defined as the lowest early or late gametocyte concentration (serially diluted cDNA from the purified stage II or stage V, respectively) detectable in all experiments that fell within the log-linear relationship of the qRT-PCR assay (Boyer et al., 2013). There was insufficient purified early gametocyte cDNA available to run a quantitative standard curve against all clinical samples, thus an ivcDNA standard curve was used. Detection of ivcDNA concentrations below the LoQ was possible in >50% of the standard curves. Although points below the LoQ were outside the log-linear relationship of the qRT-PCR, and could not be reliably quantified, they were considered to represent reliable detection because they fell within a clear amplification curve, in contrast to the cDNA-free sample (negative control) (Klymus et al., 2020).

The transcript number of the stage-specific gametocyte genes was quantified in clinical samples based on the log-linear mathematical relationship between qRT-PCR cycle threshold (C_T_) and the log10 of the concentration of the ivcDNA standard curve (supplementary Figure S1). Then, the log-linear mathematical relationship between the transcript numbers and the early gametocyte numbers in the purified stage II gametocytes culture was established (supplementary Figure S2). The number of gametocytes/mL of blood in natural infections was estimated based on the average number of transcripts obtained from known numbers of purified stage II or stage V gametocytes, assuming 100% efficiency of the processes of reverse transcription, RNA purification and RNA extraction.

### 2.7. Statistical analysis

Intra- and inter-assay coefficients of variation of the standard curves were calculated as the standard deviation of the C_T_ values divided by their mean C_T,_ and then multiplied by 100. The levels of the reference gene expression between different gametocyte stages were compared using Student’s T test. A Wilcoxon test was used to compare the relative expression of early and late gametocyte markers among stage II, stage V and asexual samples. Logistic regression was used to investigate the probability of detecting early gametocytes at variable levels of parasitaemia. Spearman’s correlation was used to assess the correlation between *pfpeg4* and *pfg27* expression in field samples. It was also used to assess the correlation between early and late gametocyte gene expression in field samples. R version 3.2.3 (2015-12-10) was used for statistical analysis.

## 3. Results

### 3.1. *In silico* analysis of gametocyte markers and the reference gene

To identify putative early-stage gametocyte markers, we searched published *P. falciparum* stage-specific gametocyte transcription analyses and transcriptome data (Bruce et al., 1994, Lobo et al., 1994, Furuya et al., 2005, Silvestrini et al., 2005, Farid et al., 2017, Eksi et al., 2012, PlasmoDB.org, 2014). Candidate genes were prioritised by comparing the transcript levels of fragments per kilobase of exon model per million mapped reads (FPKM) as described (Lopez-Barragan et al., 2011). The genes *pfpeg4, pfg27, pfge1, pfge3* and *pfgexp5* were selected for evaluation in Taqman qRT-PCR assays. The gene *pfs16* was used as a pan-gametocyte stage marker, since Pfs16 protein expression starts 24-30h post-invasion of a sexually-committed merozoite and continues throughout gametocyte development (Bruce et al., 1994). *P. falciparum* transcription data available in PlasmoDB were also searched to identify a candidate reference gene, and *pfs40S* was selected as it constitutively expressed, at the same level, in asexual parasites and gametocytes and could be used to estimate the relative expression of early gametocyte candidate genes.

### 3.2. Efficiency, reproducibility and sensitivity of qRT-PCR assay

The efficiency, sensitivity and reproducibility of the early-stage gametocyte qRT-PCR assays was assessed using ivcDNA of *pfpeg4, pfg27, pfge1, pfge3* and *pfs16*, and a late-stage gametocyte culture was used to test the *pfs25* qRT-PCR assay. In 4-6 independent experiments, the average qRT-PCR efficiency for the genes ranged between 85.6% and 92.2% and the maximum intra- and inter-assay coefficient of variation (CV) was 0.9% - 6.4% and 1.9% - 6.6% (Table S3 and Figure S1). In all cases, the low CV (<10%) demonstrates the high reproducibility of the assays. The LoQ of the *pfpeg4, pfg27, pfge1, pfge3, pfs16* and *pfs25* qPCR assays was 0.14, 0.28, 0.2, 0.14, 0.68 and 0.15 gametocyte/µL of cDNA, which is equivalent to 0.56, 1.12, 0.8, 0.56, 2.72 and 0.60 gametocytes per qRT-PCR assay, respectively.

### 3.3. Validation of early markers in purified stage II and V gametocytes

To establish that the expression level of the reference *pf40S* is similar across early and late gametocytes, qRT-PCR was performed. The reference gene, *pf40S*, showed similar expression levels (t-test T= 1.25, P= 0.28) in samples containing equal numbers of purified stage II (mean C_T_= 38.73, SD= 0.67, in 4 technical replicates of 1 sample) and stage V gametocytes (mean C_T_ = 37.60, SD= 1.43, in 4 technical replicates of 1 sample).

The stage specificity of the early gametocyte genes was assessed by quantifying their expression in comparison to the reference gene *pf40S* in three samples of *P. falciparum* culture: (i) mixed asexual stages (ii) purified stage II gametocytes and (iii) purified stage V gametocytes. The relative expression of *pfpeg4, pfg27, pfge1, pfge3* and *pfgexp5* was examined in purified stage II, stage V gametocytes (4-6 technical replicates each), and in mixed asexual stages (3-4 technical replicates). Significantly higher expression of *pfpeg4* (∼24-fold) and *pfg27* (∼43-fold) was observed in stage II compared to stage V gametocytes (Wilcoxon signed-rank test, *pfpeg4*; P=7.9×10^−3^; *pfg27*, P=0.01), with negligible expression in asexual stages (Figure 1). *Pfge1* and *pfge3* were expressed at higher levels in stage II compared to stage V, but their relative expression in stage II was considerably lower than *pfpeg4* (125-fold and 187-fold lower, respectively) and *pfg27* (50-fold and 75-fold lower, respectively). In contrast, *pfgexp5* was highly expressed in stage II gametocytes compared to asexual stages, but at higher levels in stage V gametocytes (Wilcoxon signed-rank test, P= 0.02). As expected, a significantly higher relative expression of *pfs25* (∼11-fold) was observed in stage V compared to stage II gametocytes (Wilcoxon signed-rank test, P = 7.9×10^−3^, Figure 1). Therefore, *pfpeg4* and *pfg27* were selected as early gametocyte markers for further validation in clinical samples.

**Figure 1:**
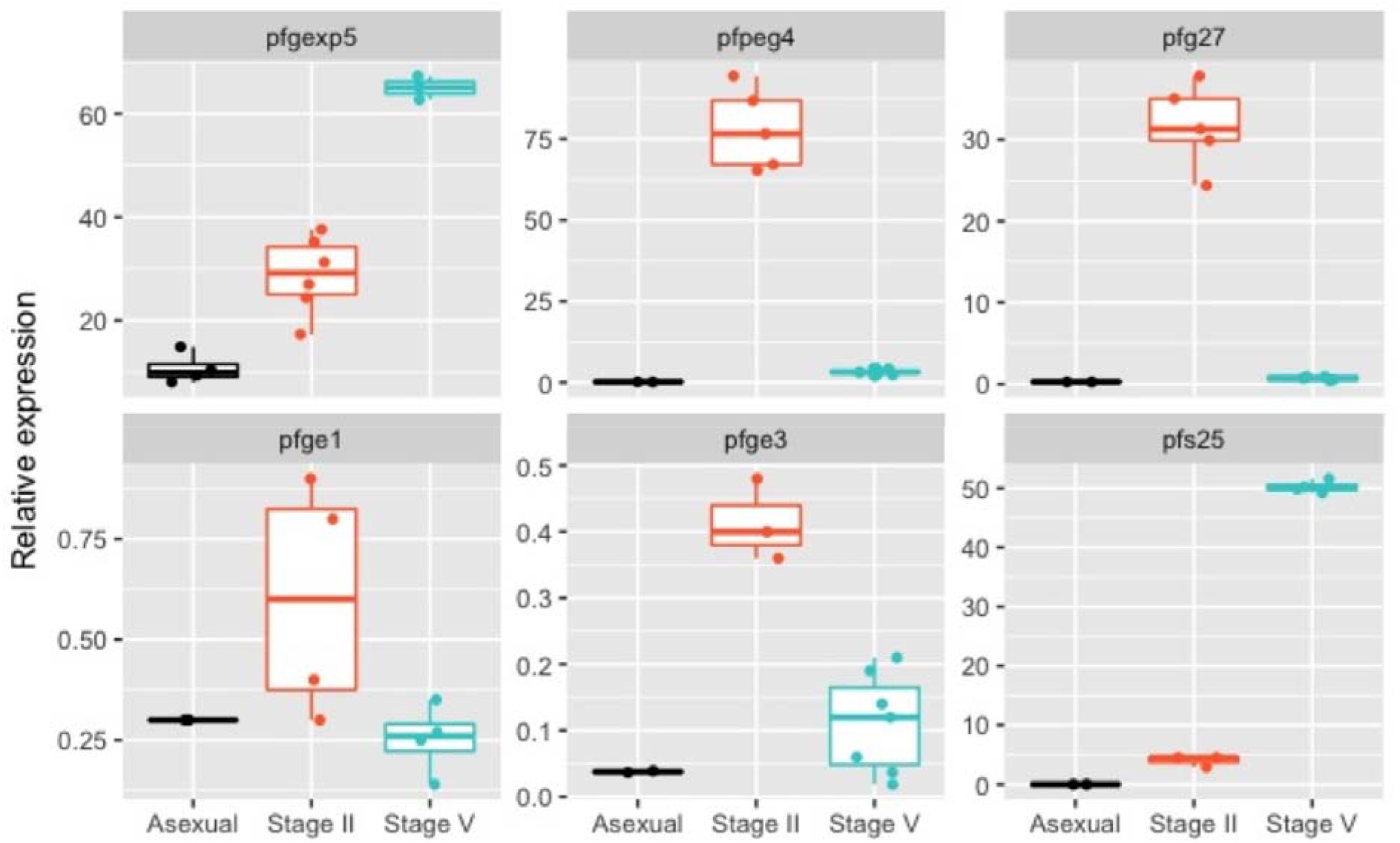
Validation of early gametocyte markers. Figure shows fold change (y axis) in expression of early (*pfpeg4, pfg27, pfge1, pfge3* and *pfgexp5*) and late (*pfs25*) markers relative to the reference gene (*pf40S*) expression in parasites obtained from *in vitro* culture at different parasite developmental stages. Points represents actual data points, boxplots represent median, first and third quartiles. Error bars represent the minimum and the maximum fold change value.

### 3.4. Detection of early and late gametocytes in natural *P. falciparum* infections

Two hundred and fifty samples from patients with natural *P. falciparum* infections (Ali et al., 2006) were used to investigate the specificity of the early gametocyte assays, across a range of parasitaemias, and in the presence of late gametocytes. Of the 250 samples, 79.2% (n=198) were positive for *pfs16*, indicating the presence of gametocytes (supplementary Table S4). Of those 198 samples, 25 (12.6%) were positive for *pfpeg4* and/or *pfg27* but not for *pfs25*, indicating the presence of early gametocytes only, and 30 (15.2%) were positive for *pfs25* and not *pfpeg4* and *pfg27*, indicating the presence of only late gametocytes (Figure 2A). The majority of the samples contained mixed early and late gametocyte stages (n = 141, 71.2%). However, for 2 (1%) samples out of the 198 that were *pfs16-*positive no transcripts for either early or late gametocyte genes were detected.

**Figure 2:**
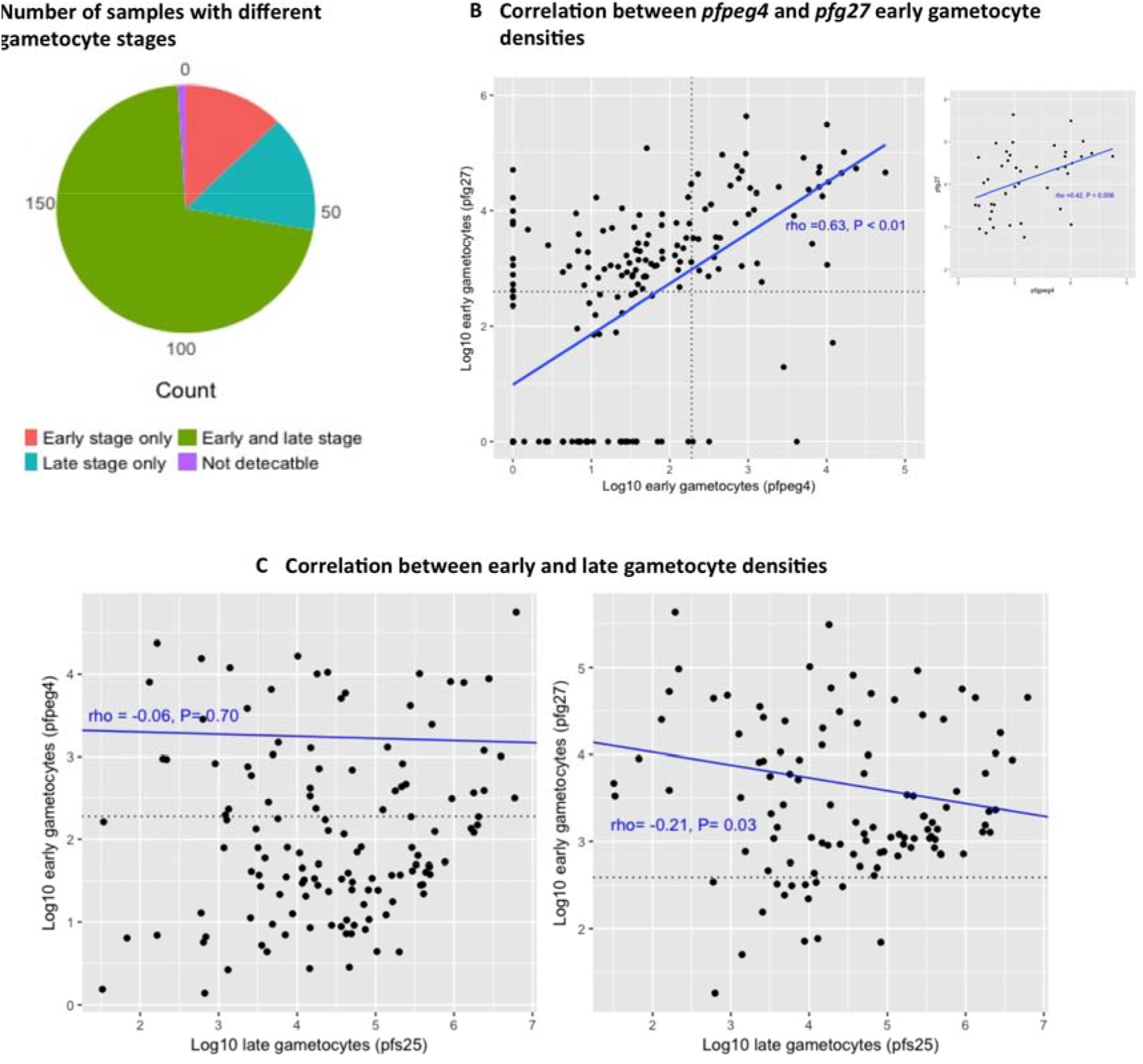
Early gametocytes in natural infections. (**A**) Number of samples with different gametocyte stages in field samples (n= 198). Early gametocyte detection is based on both *pfpeg4* and *pfg27* expression. (**B**) Correlation between log10 early gametocyte densities as deteted by *pfpeg4* and *pfg27*. The vertical and horizontal dotted lines represent the limit of quantification (LoQ) for *pfpeg4* (equivalent to 190 gametocytes/ml) and *pfg27* (equivalent to 390 gametocytes/ml), respectively. The larger panel shows correlation in all samples and the smaller panel shows correlations in samples where densities were above the LoQ (represents the right upper square on the larger panel). (**C**) Correlation between log10 early (*pfpeg4* or *pfg27*) and late stage gametocytes/ ml of blood among samples with mixed stages. The figure shows that early stage gametocytes densities are independent, particularly for *pfpeg4*, to the density of late stage gametocytes. The dotted line in both panels represents the limit of quantification. Correlation line, Spearman correlation coefficient (rho) and P value were calculated for points above the limit of quantification. Points with lower density (below LoQ line) are robustly detectable however with higher uncertainty around the estimated quantities.

The *pfpeg4* and *pfg27* qRT-PCR assays detected early gametocytes in patients with a wide range of total parasitaemia, 79 to 2.6×10^8^ parasites/mL of blood as determined by 18s rRNA qPCR. The presence of early stage gametocytes was associated with parasitaemia: for each 10-fold increase in parasite density the probability of detecting early gametocytes increased by a factor of 2.22 (95% CI= 1.63 – 3.01) for *pfpeg4* and 2.12 (95% CI= 1.67 – 2.70) for *pfg27*. Of note, >80% of samples containing early gametocytes were from patients with parasite density of >10^4^ parasites/mL of blood Figure S3.

The *pfpeg4* and *pfg27* qRT-PCR assays detected variable densities of early gametocytes, with a range of 1 to 5.6×10^4^ and 18.4 to 4.3×10^5^ early gametocytes/mL of blood, respectively. However, the densities of 104 and 19 samples were below LoQ for *pfpeg4* and *pfg27*, respectively (Section 2.6.4). The LoQ of the assays was estimated as 0.14 and 0.28 early gametocyte/µL of cDNA for *pfpeg4* and *pfg27*, equivalent to 190 and 390 early gametocytes/mL of blood, respectively, assuming 100% efficiency of the RNA extraction, purification and cDNA processes. A moderate correlation was seen between the early gametocyte densities quantified by *pfpeg4* and *pfg27* qRT-PCR assays (Spearman’s correlation coefficient (rho) =0.63, P < 0.01, Figure 2B). However, where early gametocyte densities were above the LoQ, this correlation was lower (rho =0.40, P < 0.008, Figure 2B).

Among samples containing a mix of early and late gametocytes and where early gametocyte densities were above the LoQ, early gametocyte densities did not correlate (*pfpeg4*: rho = 0.06, P= 0.70) or weakly correlated (*pfg27*: rho= −0.21, P= 0.03) with densities of late stage gametocytes and detected by *pfs25* (Figure 2C).

### 3.5. Detection of early gametocytes in experimentally infected volunteers

The specificity of the *pfg27* and *pfpeg4* assays was further tested in two volunteers experimentally infected with *P. falciparum* (Farid et al., 2017, Pasay et al., 2016). The parasite density ranged between 1.3×10^3^ – 1.6×10^5^ (median 4.4×10^4^) parasites/mL blood in volunteer S035 and 3.3×10^5^ – 8.4×10^2^ (median 5.4×10^4^) parasites/mL blood in volunteer S042 (Figure 3) over days 7 to 10 post-infection. *pfpeg4* and *pfg27* transcripts were detectable at an early stage of the infection, day 8 and day 9 post infection (Figure 3). Early gametocyte density (as identified by the *pfpeg4* and *pfg27* assays) fluctuated at low levels, often below the LoQ and with overlapping 95% CI throughout the follow up time points (*pfpeg4* range: 106 – 850; *pfg27* range: 113 – 371 early gametocytes /mL of blood). However, late gametocytes (80 to 247 gametocytes /mL) were detected below the LoQ (using the *pfs25* assay) in 4 and 5 out of the 10 time points tested in volunteer S035 and volunteer S042, respectively. The detection of late gametocytes was associated with wide 95% CI often crossing the zero due to detection occurring in only one of the sample duplicates (Figure 3).

**Figure 3:**
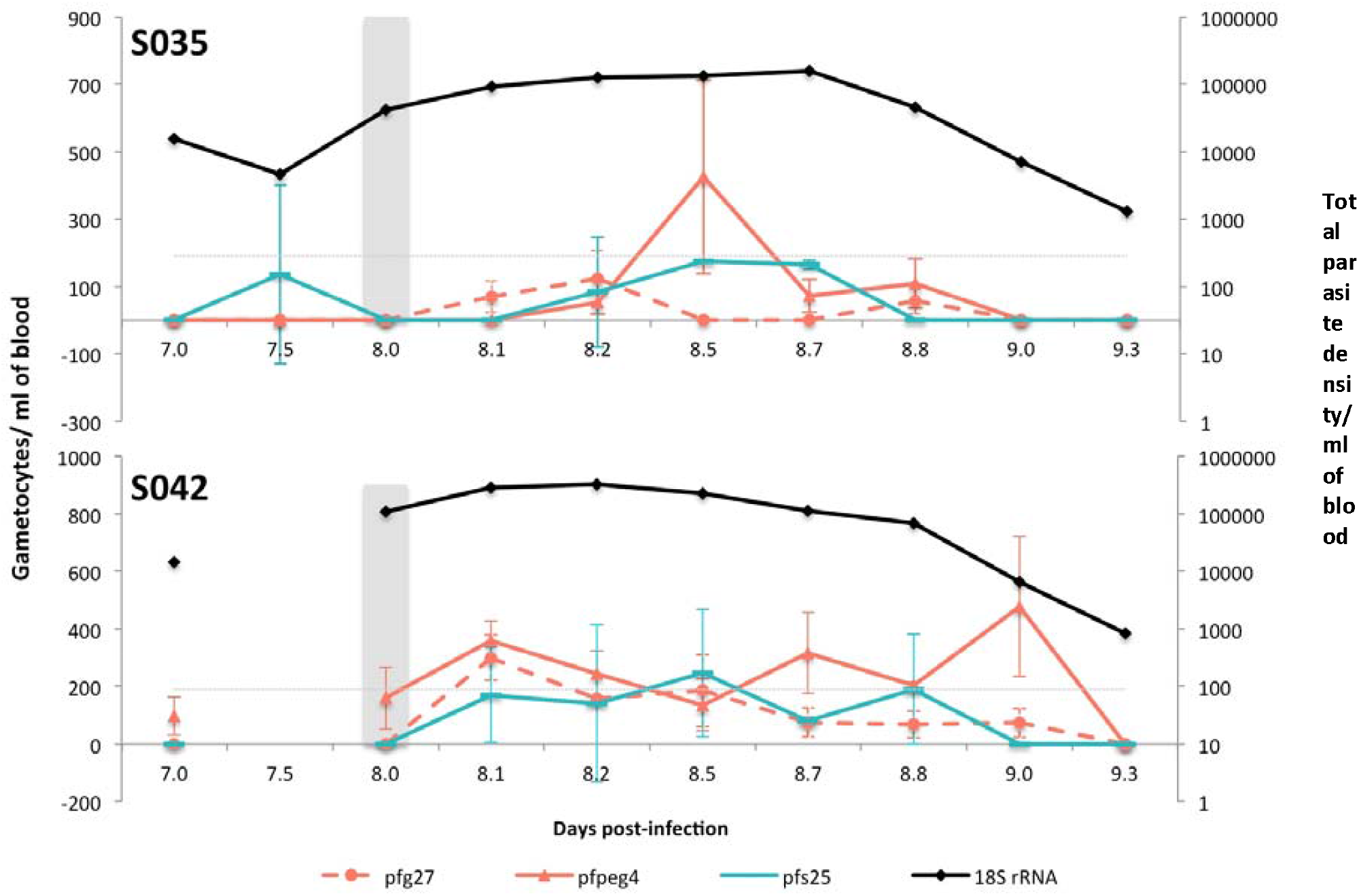
Detection of early gametocytes *in vivo* in human experimental infections. Left Y axis represents number of gametocytes/ml of blood, quantified by *pfpeg4* or *pfg27* RT-qPCR. On the right Y axis plotted total parasite density as quantified by *18S rRNA* qPCR. Horizontal grey dotted line represents limit of quantification of early gametocytes and reads to the left Y axis. Data shown is from 2 volunteers (S035 and S042) during follow up time-points between D07 and D09 post-infection. The x-axis represents day post-infection and digits after the decimal points are fractions of that day. Volunteers were treated with piperaquine at D08 (grey shaded column). Data is provided in supplementary file (Table S5).

## Discussion

The present study describes sensitive qRT-PCR assay for detection and quantification of early-stage gametocytes of *P. falciparum*. We initially examined the relative expression of five early gametocyte markers (*pfpeg4, pfg27, pfge1, pfge3* and *pfgexp5*) in purified early (stage II) and late (stage V) gametocyte and demonstrated that *pfpeg4* and *pfg27* are expressed predominantly in the early stages. In contrast, *pfgexp5* is expressed at high levels in both stage II and stage V gametocytes, and therefore is unable to distinguish between early- and late-stage gametocytes. Transcript quantification of gametocyte genes was estimated relative to that of the reference gene *pf40S*, which is expressed at the same level in stage II and V gametocytes of *P. falciparum*. Thus, variations in the amount of the starting transcripts were controlled for. The sensitivity of the *pfpeg4* and *pfg27* qRT-PCR assays were equivalent to detection thresholds of 190 and 390 gametocytes per mL of blood, respectively.

The specificity of the *pfpeg4 and pfg27* qRT-PCR assays was evident by analysis of 198 field *P. falciparum* isolates from eastern Sudan (Ali et al., 2006). The *pfpeg4* and/or *pfg27* transcripts were detected in 25 (12.6%) isolates in the absence of the *pfs25* transcripts, suggesting the presence of a cohort of circulating early gametocytes. Conversely, *pfs25* transcripts were detected in 30 (15.2%) isolates in the absence of *pfpeg4* and *pfg27* transcripts, indicating the presence of a circulating population comprising only late gametocytes (Figure 2A). Evidence of circulating early stage *P. falciparum* gametocytes in natural infections is scarce (Aguilar et al., 2014, Pelle et al., 2015), but has been documented in cases of exceedingly high parasite burdens, when asexual stages that usually sequester are also visible in circulation, as well as in splenectomized infected individuals (Bachmann et al., 2009).

Recent transcriptome analysis identified a number of early gametocyte-specific transcripts from sexually-committed ring stages (Poran et al., 2017), which were also found in peripheral blood samples in natural infections (Usui et al., 2019). The authors reported a high proportion of infections with circulating sexually-committed ring stage (76%), but the ratio of gametocytes to sexually committed rings (*in vitro*) varied dramatically, ranging from 78% to absent. This pattern is consistent with the pattern of detection of early gametocytes using *pfpeg4* and/or *pfg27* assays in our field setting in eastern Sudan. In this setting, 25 (12,6%) of 198 gametocyte-producing *P. falciparum* isolates were positive for *pfpeg4* and/or *pfg27*, but not for *pfs25*, indicating the presence of early gametocytes alone. Nonetheless, the majority of the samples (n = 141, 71.2%) contained transcripts of early (*pfpeg4* and/or *pfg27*) and late gametocyte stages (*pfs25*). These data suggest that, in addition to sexually committed ring stages, slightly older stage I gametocytes may also be found in the peripheral circulation. Further longitudinal analysis of asymptomatic infections, in the absence of re-infection in a highly seasonal endemic setting, such as that in eastern Sudan (Abdel-Wahab et al., 2002) will elucidate epidemiological factors that drive gametocytogenesis. A previous longitudinal survey of asymptomatic infection in this region demonstrated an increase in *pfs25* transcripts, indicative of mature gametocytes, following resurgence of mosquitoes at the start of the rainy season. However, no corresponding increase in the density or prevalence of total parasites or gametocytes was seen (Gadalla et al., 2016). Analysis of early markers would verify the hypothesis that *P. falciparum* may respond to environmental cues, such as mosquito biting, to modulate its transmission strategy (Reece et al., 2010).

The robustness of the *pfpeg4* and *pfg27* assays is demonstrated by the ability to detect transcripts at a wide range of parasitaemia in field samples, from very high (2.6×10^8^ parasites/mL blood), always associated with clinical presentation, to sub-microscopic levels (79 parasites/mL blood), often present in asymptomatic infections (Rogier et al., 1996, Roucher et al., 2012, Gadalla et al., 2016). The detectability of *pfpeg4* and/or *pfg27* transcripts is strongly linked to the level of total parasitaemia, in line with the performance of other qRT-PCR assays for gametocyte quantification; *pfgexp5* (Farid et al., 2017), *pfs230p* (Schneider et al., 2015) and the *pfs25* (Tadesse et al., 2017).

The specificity of the *pfpeg4* and *pfg27* qRT-PCR assays was further tested by analysis of samples from volunteers experimentally infected with *P. falciparum*, taken at D07 - D09 post-infection, when only early but not late gametocyte stages are present. The volume of the initial inoculum was 1,800 parasites; equivalent to approximately 1/10^−14^ parasite/ mL, precluding the detection of gametocytes at early time points. Transcripts of *pfpeg4* and *pfg27* were detected at significant levels (above the LoQ) at several time points between D7 and D9 (Figure 3), demonstrating the reliability of the assay, even at a low density of circulating early gametocytes at this early stage of infection. In contrast, *pfs25* transcripts were detected, but below the reliable limit of quantification (LoQ), at only two time points in each volunteer, with a wide 95% CI of replica including zero (Figure 3).

In summary, both *pfpeg4* and *pfg27* qRT-PCR assays are specific and sensitive, and can quantify early *P. falciparum* gametocytes, as low as 190 and 390 gametocytes per mL of whole blood, respectively. The late-stage specificity (*pfs25* assay), currently used for field surveys, quantifies late gametocytes (Babiker et al., 1999). These early stage-specific qRT-PCR assays will facilitate the study of epidemiological factors that influence transmission, such as the impact of drugs and the multiplicity of infection, as well as climatic variables that govern fluctuations of the mosquito vector. A better understanding of the gametocyte reservoir in natural infections is essential for novel approach for malaria elimination strategies and for the assessment of transmission blocking strategies.

## Data Availability

Data is available as supplementary file.

## Acknowledgments

We thank Department of Biochemistry staff, College of Medicine and Health Sciences, Sultan Qaboos University for support. Amal Gadalla was supported by SQU scholarship and International Atomic Energy Agency (IAEA).

