## Supplementary file for "Real-time PCR assays for detection and quantification of early *P. falciparum* gametocyte stages"

**Table S1: Genes names and Accession Numbers**

| Gene name | Annotation | Old ID | Current Accession # |
|---|---|---|---|
| *18s rRNA* | 18S ribosomal RNA | -<br>MAL11_rRNA<br>- | PF3D7_0112300<br>PF3D7_1148600<br>PF3D7_1371000 |
| *pfs16* | Sexual stage-specific protein precursor | MAL4P1.61,<br>PFD0310w | PF3D7_0406200 |
| *pfgexp5* | Gametocyte exported protein 5 | - | PF3D7_0936600 |
| *pfpeg4* | Early transcribed membrane protein 10.3 (ETRAMP10.3) | PF10_0164 | PF3D7_1016900 |
| *pfg27* | Gametocyte specific protein 27/25 | PF13_0011 | PF3D7_1302100 |
| *pfge1* | Gametocytogenesis early gene 1 | PF14_0744 | PF3D7_1477300 |
| *pfge2* | Gametocytogenesis early gene 2 | PF14_0745 | PF3D7_1477400 |
| *pfge3* | Gametocytogenesis early gene 3 | PF14_0748 | PF3D7_1477700 |
| *pfs25* | 25 kDa ookinete surface antigen precursor | PF10_0303 | PF3D7_1031000 |
| *Pf40S* | 40S ribosomal protein S3 | PF14_0627 | PF3D7_1465900 |

**Table S2: Primers and probes sequencing.** Primers and TaqMan dual labeled probes used for quantification of total parasite density, early gametocytes and late gametocytes. Fw = Forward and Rv = reverse, dual labeling in probes sequence indicated in bold, FAM = 6-carboxyfluorescein).

| **Oligo name** | **Oligonucleotide sequence (5` to 3`) and label** |
|---|---|
| pfs16 Qt.Fw | CCTTCAACTTTGCAAACCC |
| pfs16 Qt.Rv | GCTAGCTGAGTTTCTAAAGGCAT |
| pfs16 Qt.Pr | **Fam**-TGCCTCTCTTCATGCTGTTGGACC-**Tamra** |
| pfg10.164 QtFw | GCGTTGAAGGATATCGACAG |
| pfg10.164 QtRv | CCTAAACTTCCAAATAAACCACCT |
| pfg10.164 QtPr | **Fam**-TGCTGCTGTTGCTTTGGCTATAACCTT- **Tamra** |
| SP6_0164Fw1 | GACACTATAGGTTTCTAGGCATACCGTTC |
| SP6_0164Fw2 | GATTTAGGTGACACTATAGGTTTCTAGGCATACCGTTC |
| 0164Rv | AGAGTCGGATCCATCATTCTC |
| pfg27 QtFw | GCGTATCATGAACGACAAGAA |
| pfg27 QtRv | ACGGGTAAAGCAGGTATTGG |
| pfg27 QtPr | JOE-TGACCAATATAAGGACGCAGCAGCA-Tamra |
| T7-Pfg27Fw1 | ACTATAGGGTTAGTATCAGGAGATATGTTCG |
| T7-Pfg27Fw2 | TAATACGACTCACTATAGGGTTAGTATCAGGAGATATGTTCG |
| Pfg27Rv | GTTGTGATGTGGTTCATCAGGTG |
| GEXPFWD | CTTCTTGTTCGAGATTATCCCT |
| GEXPREV | GGAGTCTACTAATTCAGACAGC |
| GEXPPROBE | **JOE** -TGTAATGTAGTAGAAGGTACCATTGGTCA-**Tamra** |
| pfg14.744 QtFw (*Pfge1*) | TAATGGCTCTGTTGACGGAA |
| pfg14.744 QtRv (*Pfge1*) | TGAAGAAGAATAATTTGCAGAAGC |
| pfg14.744 QtPr (*Pfge1*) | **JOE** -TCAGAGAGGCTTCTCGACTTCCTCG-**Tamra** |
| pfg14.748 QtFw (*Pfge2*) | TCACATAATGAATTCAAGGGTAGTT |
| pfg14.748 QtRv (*Pfge2*) | TCTTCAGCTTTATCAGAATTCCC |
| pfg14.748 QtPr (*Pfge2*) | **Fam**-CTCTGATTTGGCCACACTGCTCTAGG-**Tamra** |
| pfs25 Qt.Fw | GACTGTAAATAAACCATGTGGAGATT |
| pfs25 Qt.Rv | CCGTTACCACAAGTTACATTCTTAC |
| pfs25 Qt.Pr | **FAM**- TGGAAATCCCGTTTCATACGCTTGT-T**amra** |
| Pf40s.Qt.Fw | TGCTGAAAGAGTAGAACATAGAGGA |
| Pf40s.Qt.Rv | TCCTTTAGCACCAGATTCCA |
| Pf40s.Qt.Pr | **FAM**- TCGGCTTGTGCCATTGCACA-T**amra** |

**Table S3.** Average RT-qPCR efficiency and maximum intra- and inter-assay coefficient of variation (4-6 experiments) for the genes *pfpeg4*, *pfg27*, *pfge1*, *pfge3*, *pfs16*, *pfs25*.

| | Average RT-qPCR efficiency % (SD) | intra-assay coefficient of variation % | Inter-assay coefficient of variation % |
|---|---|---|---|
| *pfpeg4* | 88.3 (13.3) | 4.2 | 2.7 |
| *pfg27* | 85.6 (4.9) | 2.4 | 4.0 |
| *pfge1* | 92.2 (12.8) | 6.4 | 5.9 |
| *pfge3* | 81.3 (7.15) | 2.3 | 6.6 |
| *pfs16* | 90.7 (11.9) | 0.9 | 1.9 |
| *pfs25* | 88.2 (0.57) | 1.2 | 3.8 |

**Table S4: Raw data for field samples**

| PID | Total parasite | Q.Total parasite | Any Gametocyte | Early (pfpeg4) | Q.Early (pfpeg4) | Early (pfg27) | Q. Early (pfg27) | Late (pfs25) | Q.Late (pfs25) | Gametocytes profile |
|---|---|---|---|---|---|---|---|---|---|---|
| E01/0 | Yes | 22981726.6 | Yes | Yes | 10086.3 | Yes | 308052.8 | Yes | 18014.5 | Early and late gametocytes |
| E01/07 | Yes | 74935.0 | No | No | 0.0 | No | 0.0 | No | 0.0 | pfs16 negative |
| E01/14 | Yes | 5188529.3 | Yes | Yes | 16632.2 | Yes | 102216.6 | Yes | 10205.0 | Early and late gametocytes |
| E01/21 | Yes | 561040.6 | Yes | Yes | 8.4 | Yes | 243.9 | Yes | 4855.9 | Early and late gametocytes |
| E03/0 | Yes | 811955.4 | Yes | Yes | 1.8 | Yes | 2492.3 | No | 0.0 | Early gametocytes only |
| E03/07 | Yes | 18696.3 | Yes | No | 0.0 | No | 0.0 | Yes | 254.7 | Late gametocytes only |
| E03/14 | Yes | 10081.8 | Yes | Yes | 32.3 | Yes | 711.8 | Yes | 36978.5 | Early and late gametocytes |
| E03/21 | Yes | 3304.0 | Yes | No | 0.0 | No | 0.0 | Yes | 1366.4 | Late gametocytes only |
| E04/0 | Yes | 15061936.5 | Yes | Yes | 929.5 | Yes | 96052.4 | Yes | 215.8 | Early and late gametocytes |
| E04/07 | Yes | 3758869.1 | Yes | Yes | 11.9 | Yes | 344.7 | Yes | 597.0 | Early and late gametocytes |
| E04/14 | Yes | 1995046.0 | Yes | Yes | 7928.9 | Yes | 44894.2 | Yes | 1335742.1 | Early and late gametocytes |
| E04/21 | Yes | 301099.7 | Yes | Yes | 11.6 | Yes | 71.7 | Yes | 8754.6 | Early and late gametocytes |
| E07/0 | Yes | 2656414.1 | Yes | Yes | 5.9 | Yes | 3887.8 | Yes | 163.6 | Early and late gametocytes |
| E07/07 | Yes | 6041387.7 | Yes | Yes | 44.1 | Yes | 436.4 | Yes | 11639.2 | Early and late gametocytes |
| E07/14 | Yes | 1017725.2 | Yes | Yes | 690.3 | Yes | 6061.5 | Yes | 50415.5 | Early and late gametocytes |
| E07/21 | Yes | 1616603.8 | Yes | Yes | 58.3 | Yes | 326.9 | Yes | 3897.1 | Early and late gametocytes |
| E08/0 | Yes | 14059118.2 | Yes | No | 0.0 | Yes | 6498.9 | No | 0.0 | Early gametocytes only |
| E08/07 | Yes | 341.1 | No | No | 0.0 | No | 0.0 | No | 0.0 | pfs16 negative |
| E08/14 | Yes | 202225.1 | Yes | Yes | 435.2 | Yes | 3350.8 | Yes | 214465.0 | Early and late gametocytes |
| E08/21 | Yes | 207369.2 | Yes | No | 0.0 | No | 0.0 | Yes | 94.5 | Late gametocytes only |
| E9/0 | Yes | 5441178.7 | Yes | Yes | 115.4 | Yes | 1665.7 | Yes | 39599.3 | Early and late gametocytes |
| E9/07 | Yes | 2401.1 | Yes | No | 0.0 | No | 0.0 | Yes | 10586.5 | Late gametocytes only |
| E9/14 | Yes | 1057.3 | No | No | 0.0 | No | 0.0 | Yes | 13302.4 | pfs16 negative |
| E9/21 | Yes | 2024.9 | No | No | 0.0 | No | 0.0 | Yes | 5295.4 | pfs16 negative |
| E11/0 | Yes | 6218954.1 | Yes | Yes | 35.0 | Yes | 371.2 | No | 0.0 | Early gametocytes only |
| E11/07 | Yes | 102429.6 | Yes | Yes | 9.7 | Yes | 69.8 | Yes | 82598.1 | Early and late gametocytes |
| E11/14 | Yes | 233765.9 | Yes | Yes | 22.5 | No | 0.0 | Yes | 25225.9 | Early and late gametocytes |
| E11/21 | Yes | 23548.4 | Yes | No | 0.0 | No | 0.0 | Yes | 5645.0 | Late gametocytes only |
| E13/0 | Yes | 2794108.4 | Yes | Yes | 0.5 | Yes | 4670.6 | Yes | 32.0 | Early and late gametocytes |

| | | | | | | | | | | |
|---|---|---|---|---|---|---|---|---|---|---|
| E13/07 | Yes | 272204.6 | Yes | Yes | 1.6 | No | 0.0 | Yes | 1314.1 | Early and late gametocytes |
| E13/14 | Yes | 227.2 | No | No | 0.0 | No | 0.0 | No | 0.0 | pfs16 negative |
| E15/0 | Yes | 92257492.2 | Yes | Yes | 38.6 | Yes | 2709.0 | No | 0.0 | Early gametocytes only |
| E15/07 | Yes | 382587.2 | Yes | Yes | 36.1 | No | 0.0 | Yes | 206261.8 | Early and late gametocytes |
| E15/14 | Yes | 11425439.5 | Yes | Yes | 62.6 | Yes | 1088.0 | Yes | 346815.7 | Early and late gametocytes |
| E15/21 | Yes | 6272318.4 | Yes | Yes | 38.2 | Yes | 519.8 | Yes | 44677.3 | Early and late gametocytes |
| E16/0 | Yes | 71021820.3 | Yes | Yes | 1273.4 | Yes | 20074.7 | Yes | 14935.2 | Early and late gametocytes |
| E17/0 | Yes | 19938337.9 | Yes | Yes | 36.0 | Yes | 2088.6 | Yes | 3269.7 | Early and late gametocytes |
| E17/07 | Yes | 5833.8 | Yes | No | 0.0 | No | 0.0 | Yes | 50760.0 | Late gametocytes only |
| E17/14 | Yes | 3986.6 | Yes | No | 0.0 | No | 0.0 | Yes | 23741.2 | Late gametocytes only |
| E17/21 | Yes | 910.5 | Yes | No | 0.0 | No | 0.0 | Yes | 5135.0 | Late gametocytes only |
| E18/0 | Yes | 25348947.3 | Yes | Yes | 8.0 | Yes | 1887.2 | No | 0.0 | Early gametocytes only |
| E18/07 | Yes | 66497.6 | Yes | Yes | 7.7 | No | 0.0 | No | 0.0 | Early gametocytes only |
| E18/14 | Yes | 13914888.7 | Yes | Yes | 23771.1 | Yes | 52983.9 | Yes | 163.6 | Early and late gametocytes |
| E18/21 | Yes | 23341.5 | Yes | No | 0.0 | No | 0.0 | Yes | 418.6 | Late gametocytes only |
| E19/0 | Yes | 413570.4 | No | No | 0.0 | No | 0.0 | No | 0.0 | pfs16 negative |
| E19/07 | Yes | 9535.0 | Yes | No | 0.0 | No | 0.0 | Yes | 33213.4 | Late gametocytes only |
| E19/21 | Yes | 61706.7 | Yes | Yes | 23.6 | No | 0.0 | Yes | 49980.6 | Early and late gametocytes |
| E20/0 | Yes | 8283514.6 | Yes | No | 0.0 | No | 0.0 | No | 0.0 | Fail to detect early or late markers |
| E20/07 | Yes | 7418296.4 | Yes | Yes | 8135.7 | Yes | 56485.3 | Yes | 902612.5 | Early and late gametocytes |
| E20/14 | Yes | 14583251.0 | Yes | Yes | 8020.2 | Yes | 25200.9 | Yes | 130.4 | Early and late gametocytes |
| E20/21 | Yes | 35074.3 | Yes | Yes | 79.8 | Yes | 1453.9 | Yes | 65675.8 | Early and late gametocytes |
| E22/0 | Yes | 3407853.8 | Yes | Yes | 5.8 | Yes | 1982.2 | No | 0.0 | Early gametocytes only |
| E22/07 | Yes | 2789.9 | Yes | Yes | 5.6 | No | 0.0 | Yes | 685.6 | Early and late gametocytes |
| E22/14 | Yes | 1122.2 | Yes | No | 0.0 | No | 0.0 | Yes | 177.6 | Late gametocytes only |
| E22/21 | Yes | 756.6 | No | No | 0.0 | No | 0.0 | No | 0.0 | pfs16 negative |
| E23/0 | Yes | 25859468.8 | Yes | Yes | 77.9 | Yes | 5598.4 | Yes | 3175.0 | Early and late gametocytes |
| E23/07 | Yes | 1315364.7 | Yes | Yes | 56.8 | Yes | 1188.1 | No | 0.0 | Early gametocytes only |
| E23/14 | Yes | 2167722.2 | Yes | Yes | 26.0 | No | 0.0 | Yes | 3432.6 | Early and late gametocytes |
| E25/0 | Yes | 23714128.9 | Yes | Yes | 170.7 | Yes | 17126.3 | Yes | 1268.8 | Early and late gametocytes |
| E25/07 | Yes | 802809.6 | Yes | Yes | 310.3 | Yes | 718.1 | Yes | 938228.6 | Early and late gametocytes |
| E25/14 | Yes | 676006.1 | Yes | Yes | 31.3 | Yes | 341.3 | Yes | 12311.8 | Early and late gametocytes |

| | | | | | | | | | | |
|---|---|---|---|---|---|---|---|---|---|---|
| E25/21 | Yes | 18198.8 | Yes | Yes | 26.9 | No | 0.0 | Yes | 18370.6 | Early and late gametocytes |
| E26/0 | Yes | 18995660.2 | Yes | Yes | 40.0 | Yes | 8406.4 | Yes | 2611.6 | Early and late gametocytes |
| E26/07 | Yes | 537.9 | No | No | 0.0 | No | 0.0 | Yes | 1093.0 | pfs16 negative |
| E26/14 | Yes | 1223.0 | Yes | No | 0.0 | Yes | 321.4 | Yes | 8960.8 | Early and late gametocytes |
| E26/21 | Yes | 728.8 | No | No | 0.0 | Yes | 592.2 | Yes | 7294.2 | pfs16 negative |
| E27/0 | Yes | 9514878.9 | Yes | Yes | 10.6 | Yes | 16943.5 | No | 0.0 | Early gametocytes only |
| E27/07 | Yes | 4003.0 | Yes | No | 0.0 | No | 0.0 | Yes | 8154.6 | Late gametocytes only |
| E27/14 | Yes | 1441.8 | Yes | No | 0.0 | Yes | 305.2 | Yes | 26825.7 | Early and late gametocytes |
| E27/21 | Yes | 612.5 | Yes | No | 0.0 | No | 0.0 | Yes | 11985.1 | Late gametocytes only |
| E28/0 | Yes | 26853002.0 | Yes | Yes | 4.2 | Yes | 1082.8 | Yes | 3548.1 | Early and late gametocytes |
| E28/07 | Yes | 1105.4 | No | No | 0.0 | No | 0.0 | Yes | 3780.7 | pfs16 negative |
| E28/14 | Yes | 150.9 | No | No | 0.0 | No | 0.0 | Yes | 1867.4 | pfs16 negative |
| E31/0 | Yes | 14779603.5 | Yes | Yes | 36.0 | Yes | 2052.3 | No | 0.0 | Early gametocytes only |
| E31/07 | Yes | 1393.5 | Yes | Yes | 0.4 | No | 0.0 | Yes | 664.7 | Early and late gametocytes |
| E31/14 | Yes | 2170.0 | Yes | No | 0.0 | No | 0.0 | Yes | 198.0 | Late gametocytes only |
| E31/21 | Yes | 211.8 | No | No | 0.0 | No | 0.0 | Yes | 32.0 | pfs16 negative |
| E32/0 | Yes | 27170927.7 | Yes | No | 0.0 | No | 0.0 | No | 0.0 | Fail to detect early or late markers |
| E32/07 | Yes | 113172.6 | Yes | Yes | 6.0 | No | 0.0 | Yes | 7077.6 | Early and late gametocytes |
| E32/14 | Yes | 5739809.6 | Yes | Yes | 126.6 | Yes | 928.6 | Yes | 24946.6 | Early and late gametocytes |
| E32/21 | Yes | 257943.4 | Yes | No | 0.0 | No | 0.0 | Yes | 598.1 | Late gametocytes only |
| E35/0 | Yes | 36096722.7 | Yes | Yes | 13.1 | Yes | 4461.3 | No | 0.0 | Early gametocytes only |
| E35/07 | Yes | 1380.4 | Yes | No | 0.0 | No | 0.0 | Yes | 3780.7 | Late gametocytes only |
| E35/14 | Yes | 1048.2 | Yes | Yes | 1.7 | No | 0.0 | Yes | 14455.9 | Early and late gametocytes |
| E35/21 | Yes | 395.2 | No | No | 0.0 | No | 0.0 | No | 0.0 | pfs16 negative |
| E36/0 | Yes | 15004209.0 | Yes | Yes | 15529.9 | Yes | 44185.2 | Yes | 602.3 | Early and late gametocytes |
| E36/07 | Yes | 773661.3 | Yes | No | 0.0 | Yes | 316.7 | No | 0.0 | Early gametocytes only |
| E36/14 | Yes | 5313979.0 | Yes | Yes | 236.3 | Yes | 905.4 | Yes | 17425.0 | Early and late gametocytes |
| E36/21 | Yes | 695887.3 | Yes | Yes | 4.7 | No | 0.0 | Yes | 640.2 | Early and late gametocytes |
| E37/0 | Yes | 11546363.3 | Yes | Yes | 176.7 | Yes | 5960.5 | Yes | 5694.5 | Early and late gametocytes |
| E37/07 | Yes | 26738.5 | No | No | 0.0 | No | 0.0 | No | 0.0 | pfs16 negative |
| E37/14 | Yes | 146800.3 | No | No | 0.0 | No | 0.0 | Yes | 4601.9 | pfs16 negative |
| E37/21 | Yes | 11271.3 | Yes | Yes | 9.6 | No | 0.0 | Yes | 42812.4 | Early and late gametocytes |

| | | | | | | | | | | |
|---|---|---|---|---|---|---|---|---|---|---|
| E38/0 | Yes | 69345468.8 | Yes | Yes | 718.3 | Yes | 58075.2 | Yes | 19060.9 | Early and late gametocytes |
| E38/07 | Yes | 177105.3 | Yes | Yes | 6.2 | No | 0.0 | Yes | 42293.2 | Early and late gametocytes |
| E38/14 | Yes | 74797.1 | Yes | Yes | 123.5 | Yes | 2473.9 | Yes | 568738.6 | Early and late gametocytes |
| E38/21 | Yes | 23878785.2 | Yes | Yes | 227.6 | Yes | 42464.7 | Yes | 123877.0 | Early and late gametocytes |
| E39/0 | Yes | 94583562.5 | Yes | Yes | 6542.7 | Yes | 2642.3 | Yes | 4689.4 | Early and late gametocytes |
| E39/07 | Yes | 14130.7 | Yes | Yes | 48.3 | Yes | 1375.0 | Yes | 328319.0 | Early and late gametocytes |
| E39/14 | Yes | 11929.5 | Yes | Yes | 15.3 | No | 0.0 | Yes | 70863.2 | Early and late gametocytes |
| E39/21 | Yes | 9494.6 | Yes | Yes | 11.2 | Yes | 679.7 | Yes | 136285.5 | Early and late gametocytes |
| E43/0 | Yes | 20040695.3 | Yes | Yes | 78.8 | Yes | 8660.0 | Yes | 7520.7 | Early and late gametocytes |
| E43/07 | Yes | 20197.8 | Yes | Yes | 8.1 | No | 0.0 | Yes | 27570.7 | Early and late gametocytes |
| E43/14 | Yes | 2320.1 | Yes | Yes | 16.6 | Yes | 1108.8 | Yes | 165085.3 | Early and late gametocytes |
| E43/21 | Yes | 393.3 | No | No | 0.0 | No | 0.0 | No | 0.0 | pfs16 negative |
| E44/0 | Yes | 206369687.5 | Yes | Yes | 5900.0 | Yes | 22811.9 | Yes | 41237.3 | Early and late gametocytes |
| E44/07 | Yes | 509719.6 | Yes | Yes | 23.3 | No | 0.0 | Yes | 107545.6 | Early and late gametocytes |
| E44/14 | Yes | 37884140.6 | Yes | Yes | 2452.6 | Yes | 25273.2 | Yes | 522875.5 | Early and late gametocytes |
| E44/21 | Yes | 53352.7 | Yes | Yes | 1184.4 | Yes | 10338.7 | Yes | 2415962.2 | Early and late gametocytes |
| E45/0 | Yes | 4398569.3 | Yes | No | 0.0 | No | 0.0 | Yes | 2825.4 | Late gametocytes only |
| E45/07 | Yes | 6260.0 | Yes | No | 0.0 | No | 0.0 | Yes | 50564.0 | Late gametocytes only |
| E45/14 | Yes | 3535.6 | Yes | Yes | 8.1 | Yes | 1021.9 | Yes | 53351.0 | Early and late gametocytes |
| E45/21 | Yes | 2838.4 | No | No | 0.0 | No | 0.0 | Yes | 12479.3 | pfs16 negative |
| E46/0 | Yes | 8916243.2 | No | No | 0.0 | No | 0.0 | No | 0.0 | pfs16 negative |
| E46/07 | Yes | 148162.5 | Yes | Yes | 386.6 | Yes | 3447.7 | Yes | 178556.2 | Early and late gametocytes |
| E46/14 | Yes | 66629.2 | Yes | Yes | 7.5 | No | 0.0 | Yes | 14659.9 | Early and late gametocytes |
| E46/21 | Yes | 23672.6 | No | No | 0.0 | No | 0.0 | Yes | 197.2 | pfs16 negative |
| E51/0 | Yes | 20595726.6 | Yes | Yes | 759.2 | Yes | 35477.9 | Yes | 2348.8 | Early and late gametocytes |
| E51/07 | Yes | 58892.8 | Yes | Yes | 3.3 | Yes | 850.2 | Yes | 200344.2 | Early and late gametocytes |
| E51/14 | Yes | 4202.0 | Yes | No | 0.0 | No | 0.0 | Yes | 39014.1 | Late gametocytes only |
| E51/21 | Yes | 1931.2 | No | No | 0.0 | No | 0.0 | No | 0.0 | pfs16 negative |
| E52/0 | Yes | 134177.7 | Yes | Yes | 120.5 | Yes | 6104.4 | Yes | 1792258.9 | Early and late gametocytes |
| E52/07 | Yes | 9374.4 | Yes | Yes | 21.0 | Yes | 1056.7 | Yes | 408504.9 | Early and late gametocytes |
| E52/14 | Yes | 1313.9 | Yes | Yes | 35.9 | Yes | 929.3 | Yes | 160777.0 | Early and late gametocytes |
| E52/21 | Yes | 1671.4 | Yes | No | 0.0 | No | 0.0 | Yes | 41024.0 | Late gametocytes only |

| | | | | | | | | | | |
|---|---|---|---|---|---|---|---|---|---|---|
| E54/0 | Yes | 3480140.6 | Yes | Yes | 23.6 | Yes | 10980.2 | No | 0.0 | Early gametocytes only |
| E54/07 | Yes | 12094.4 | No | No | 0.0 | No | 0.0 | No | 0.0 | pfs16 negative |
| E54/14 | Yes | 14068.4 | Yes | Yes | 47.5 | Yes | 721.1 | Yes | 479610.4 | Early and late gametocytes |
| E54/21 | Yes | 913.2 | Yes | No | 0.0 | No | 0.0 | Yes | 15036.1 | Late gametocytes only |
| E55/0 | Yes | 14256019.5 | No | No | 0.0 | No | 0.0 | No | 0.0 | pfs16 negative |
| E55/07 | Yes | 397.3 | No | No | 0.0 | No | 0.0 | No | 0.0 | pfs16 negative |
| E55/14 | Yes | 753.0 | No | No | 0.0 | No | 0.0 | No | 0.0 | pfs16 negative |
| E56/0 | Yes | 16307435.5 | Yes | Yes | 5.4 | Yes | 8946.1 | Yes | 67.0 | Early and late gametocytes |
| E56/07 | Yes | 581595.8 | Yes | Yes | 56101.2 | Yes | 45166.3 | Yes | 6202978.6 | Early and late gametocytes |
| E56/14 | Yes | 135019.7 | Yes | Yes | 8833.7 | Yes | 17747.0 | Yes | 2786060.3 | Early and late gametocytes |
| E56/21 | Yes | 20160.6 | Yes | Yes | 32.6 | No | 0.0 | Yes | 16666.4 | Early and late gametocytes |
| E57/0 | Yes | 69551710.9 | Yes | Yes | 592.5 | Yes | 26682.7 | Yes | 2609.7 | Early and late gametocytes |
| E57/07 | Yes | 328.5 | Yes | Yes | 17.3 | Yes | 4976.0 | No | 0.0 | Early gametocytes only |
| E57/14 | Yes | 3608.0 | No | No | 0.0 | Yes | 809.9 | No | 0.0 | pfs16 negative |
| E57/21 | Yes | 908.9 | Yes | No | 0.0 | Yes | 766.3 | Yes | 1535.4 | Early and late gametocytes |
| E59/0 | Yes | 6954930.7 | Yes | Yes | 186.5 | Yes | 28401.1 | Yes | 283199.5 | Early and late gametocytes |
| E59/07 | Yes | 19932.9 | No | No | 0.0 | No | 0.0 | Yes | 1794.5 | pfs16 negative |
| E59/14 | Yes | 1559.1 | Yes | Yes | 29.5 | Yes | 1228.1 | Yes | 50540.8 | Early and late gametocytes |
| E59/21 | Yes | 2698.0 | Yes | No | 0.0 | Yes | 221.3 | Yes | 9796.4 | Early and late gametocytes |
| E60/0 | Yes | 54627746.1 | Yes | Yes | 827.7 | Yes | 47934.1 | Yes | 903.9 | Early and late gametocytes |
| E60/07 | Yes | 18678.6 | Yes | Yes | 20.6 | Yes | 312.7 | Yes | 6013.7 | Early and late gametocytes |
| E60/14 | Yes | 1132091.2 | Yes | Yes | 1.1 | No | 0.0 | No | 0.0 | Early gametocytes only |
| E60/21 | Yes | 1690.9 | Yes | Yes | 10.2 | Yes | 155.5 | Yes | 2550.1 | Early and late gametocytes |
| E61/0 | Yes | 13537384.8 | Yes | Yes | 465.7 | Yes | 92143.9 | Yes | 243992.7 | Early and late gametocytes |
| E61/07 | Yes | 21809.3 | Yes | Yes | 32.8 | Yes | 763.0 | Yes | 90314.8 | Early and late gametocytes |
| E61/14 | Yes | 19594.6 | Yes | Yes | 78.5 | Yes | 1953.3 | Yes | 290359.8 | Early and late gametocytes |
| E61/21 | Yes | 178769.2 | Yes | No | 0.0 | No | 0.0 | Yes | 3698.0 | Late gametocytes only |
| E63/0 | Yes | 93298984.4 | Yes | Yes | 5084.2 | Yes | 81414.3 | Yes | 36498.0 | Early and late gametocytes |
| E63/07 | Yes | 45834.2 | Yes | No | 0.0 | Yes | 521.7 | No | 0.0 | Early gametocytes only |
| E63/14 | Yes | 10597.2 | Yes | Yes | 1009.1 | Yes | 8649.5 | Yes | 3958566.7 | Early and late gametocytes |
| E63/21 | Yes | 413.4 | Yes | Yes | 23.5 | Yes | 741.7 | Yes | 80969.9 | Early and late gametocytes |
| E66/0 | Yes | 38645589.8 | Yes | Yes | 2835.5 | Yes | 18.4 | Yes | 629.4 | Early and late gametocytes |

| | | | | | | | | | | |
|---|---|---|---|---|---|---|---|---|---|---|
| E66/07 | Yes | 1511541.4 | Yes | Yes | 4156.7 | No | 0.0 | Yes | 278132.5 | Early and late gametocytes |
| E66/14 | Yes | 3536696.3 | Yes | Yes | 825.3 | Yes | 1084.0 | Yes | 220708.1 | Early and late gametocytes |
| E66/21 | Yes | 11147.0 | Yes | Yes | 171.5 | No | 0.0 | Yes | 22866.7 | Early and late gametocytes |
| E68/0 | Yes | 197907.5 | Yes | Yes | 38.9 | Yes | 1386.5 | Yes | 436387.3 | Early and late gametocytes |
| E68/07 | Yes | 148815.5 | No | No | 0.0 | No | 0.0 | No | 0.0 | pfs16 negative |
| E68/14 | Yes | 29705.4 | Yes | Yes | 40.6 | Yes | 1967.7 | Yes | 295348.9 | Early and late gametocytes |
| E68/21 | Yes | 5079.6 | Yes | No | 0.0 | Yes | 409.9 | Yes | 67946.4 | Early and late gametocytes |
| E69/0 | Yes | 2789264.2 | Yes | Yes | 13.6 | Yes | 958.1 | No | 0.0 | Early gametocytes only |
| E69/07 | Yes | 62.8 | No | No | 0.0 | No | 0.0 | No | 0.0 | pfs16 negative |
| E69/14 | Yes | 2167.0 | No | No | 0.0 | No | 0.0 | No | 0.0 | pfs16 negative |
| E70/0 | Yes | 2457231.9 | Yes | Yes | 1060.9 | Yes | 24201.1 | Yes | 4919.1 | Early and late gametocytes |
| E70/07 | Yes | 2215055.7 | Yes | No | 0.0 | No | 0.0 | Yes | 75633.7 | Late gametocytes only |
| E70/14 | Yes | 27577.8 | Yes | Yes | 366.3 | Yes | 1541.8 | Yes | 1783436.9 | Early and late gametocytes |
| E70/21 | Yes | 56413.2 | Yes | Yes | 390.4 | Yes | 2315.4 | Yes | 2418283.4 | Early and late gametocytes |
| E71/0 | Yes | 12023570.3 | Yes | Yes | 231.0 | Yes | 3197.8 | Yes | 1340.0 | Early and late gametocytes |
| E71/14 | Yes | 157.1 | No | No | 0.0 | No | 0.0 | Yes | 2415.5 | pfs16 negative |
| E71/21 | Yes | 339.7 | No | No | 0.0 | No | 0.0 | Yes | 3470.2 | pfs16 negative |
| E72/0 | Yes | 12060939.5 | Yes | Yes | 11961.0 | Yes | 50.4 | Yes | 1394.2 | Early and late gametocytes |
| E72/07 | Yes | 2287.5 | Yes | Yes | 77.8 | No | 0.0 | Yes | 1166.3 | Early and late gametocytes |
| E72/14 | Yes | 295.3 | No | No | 0.0 | No | 0.0 | Yes | 105.8 | pfs16 negative |
| E72/21 | Yes | 455.3 | No | Yes | 17.7 | No | 0.0 | Yes | 18.9 | pfs16 negative |
| E73/0 | Yes | 927685.4 | Yes | Yes | 132.3 | Yes | 465.4 | Yes | 3000.4 | Early and late gametocytes |
| E73/07 | Yes | 1318.0 | Yes | No | 0.0 | No | 0.0 | Yes | 33899.6 | Late gametocytes only |
| E73/21 | Yes | 231.7 | No | No | 0.0 | No | 0.0 | No | 0.0 | pfs16 negative |
| E75/0 | Yes | 19933832.0 | Yes | Yes | 147.9 | Yes | 2213.9 | Yes | 1990302.2 | Early and late gametocytes |
| E75/07 | Yes | 189805.1 | Yes | Yes | 186.8 | Yes | 1273.6 | Yes | 2056259.0 | Early and late gametocytes |
| E75/14 | Yes | 109240.4 | Yes | Yes | 134.2 | Yes | 1284.4 | Yes | 1651845.3 | Early and late gametocytes |
| E75/21 | Yes | 44827.7 | Yes | Yes | 45.5 | Yes | 703.2 | Yes | 485451.5 | Early and late gametocytes |
| E78/0 | Yes | 2572113.0 | Yes | Yes | 281.6 | Yes | 10647.8 | Yes | 4311.9 | Early and late gametocytes |
| E78/07 | Yes | 832.4 | Yes | Yes | 6.2 | No | 0.0 | Yes | 49096.3 | Early and late gametocytes |
| E78/14 | Yes | 499.8 | Yes | No | 0.0 | No | 0.0 | Yes | 23957.2 | Late gametocytes only |
| E78/21 | Yes | 863917.1 | Yes | Yes | 67.1 | Yes | 1108.9 | Yes | 10726.6 | Early and late gametocytes |

| | | | | | | | | | | |
|---|---|---|---|---|---|---|---|---|---|---|
| E79/0 | Yes | 1479102.4 | Yes | Yes | 7.1 | Yes | 502.0 | Yes | 74124.6 | Early and late gametocytes |
| E79/07 | Yes | 768.9 | Yes | Yes | 1.8 | No | 0.0 | Yes | 46264.8 | Early and late gametocytes |
| E79/21 | Yes | 54.2 | No | No | 0.0 | No | 0.0 | No | 0.0 | pfs16 negative |
| E80/0 | Yes | 1417797.4 | Yes | Yes | 19.6 | Yes | 77.0 | Yes | 12920.4 | Early and late gametocytes |
| E80/07 | Yes | 1070.7 | No | No | 0.0 | No | 0.0 | No | 0.0 | pfs16 negative |
| E80/14 | Yes | 199.1 | No | No | 0.0 | No | 0.0 | Yes | 1803.1 | pfs16 negative |
| E80/21 | Yes | 270.5 | No | No | 0.0 | No | 0.0 | Yes | 2038.8 | pfs16 negative |
| E81/0 | Yes | 49606375.0 | Yes | Yes | 1485.8 | Yes | 571.9 | Yes | 5765.1 | Early and late gametocytes |
| E81/07 | Yes | 5600.6 | No | No | 0.0 | No | 0.0 | No | 0.0 | pfs16 negative |
| E81/14 | Yes | 348.3 | No | No | 0.0 | No | 0.0 | Yes | 6068.1 | pfs16 negative |
| E82/0 | Yes | 2354426.3 | Yes | Yes | 52.3 | Yes | 3784.0 | Yes | 766903.0 | Early and late gametocytes |
| E82/07 | Yes | 3258.0 | Yes | Yes | 3.4 | No | 0.0 | Yes | 104191.9 | Early and late gametocytes |
| E82/14 | Yes | 46244.0 | No | No | 0.0 | No | 0.0 | No | 0.0 | pfs16 negative |
| E82/21 | Yes | 407666.0 | Yes | Yes | 34.2 | Yes | 5143.0 | Yes | 7267.5 | Early and late gametocytes |
| E84/0 | Yes | 296350.0 | Yes | Yes | 49.8 | Yes | 119637.3 | No | 0.0 | Early gametocytes only |
| E84/14 | Yes | 18671.6 | Yes | Yes | 334.7 | Yes | 12831.2 | Yes | 14633.7 | Early and late gametocytes |
| E84/21 | Yes | 307.6 | No | No | 0.0 | No | 0.0 | Yes | 165.8 | pfs16 negative |
| E85/0 | Yes | 3181826.2 | Yes | Yes | 5.6 | Yes | 89.4 | No | 0.0 | Early gametocytes only |
| E85/14 | Yes | 1716072.3 | Yes | Yes | 27.5 | Yes | 846.4 | Yes | 399751.8 | Early and late gametocytes |
| E85/21 | Yes | 1146713.4 | Yes | Yes | 27.0 | Yes | 1654.3 | Yes | 375423.1 | Early and late gametocytes |
| E89/0 | Yes | 7940950.2 | Yes | Yes | 49.2 | Yes | 2638.7 | Yes | 18752.2 | Early and late gametocytes |
| E89/07 | Yes | 810479.7 | Yes | Yes | 316.1 | No | 0.0 | Yes | 5928659.9 | Early and late gametocytes |
| E89/14 | Yes | 155395.1 | Yes | Yes | 36.7 | No | 0.0 | Yes | 494843.7 | Early and late gametocytes |
| E89/21 | Yes | 67513.8 | Yes | No | 0.0 | No | 0.0 | Yes | 34632.7 | Late gametocytes only |
| E90/0 | Yes | 21197671.9 | Yes | Yes | 944.5 | Yes | 429353.2 | Yes | 194.4 | Early and late gametocytes |
| E90/07 | Yes | 85792.2 | Yes | Yes | 1298.6 | Yes | 1207.5 | Yes | 142418.0 | Early and late gametocytes |
| E90/14 | Yes | 16687845.7 | Yes | Yes | 418.1 | Yes | 963.9 | Yes | 14699.6 | Early and late gametocytes |
| E90/21 | Yes | 717095.8 | Yes | No | 0.0 | No | 0.0 | Yes | 194.9 | Late gametocytes only |
| E92/0 | Yes | 4030785.6 | Yes | No | 0.0 | Yes | 5801.5 | No | 0.0 | Early gametocytes only |
| E92/07 | Yes | 3935.6 | Yes | No | 0.0 | No | 0.0 | Yes | 3083.9 | Late gametocytes only |
| E92/14 | Yes | 5779.4 | Yes | No | 0.0 | Yes | 1454.4 | Yes | 3917.1 | Early and late gametocytes |
| E92/21 | Yes | 1673.5 | Yes | No | 0.0 | Yes | 24385043.1 | Yes | 7523.3 | Early and late gametocytes |

| | | | | | | | | | | |
|---|---|---|---|---|---|---|---|---|---|---|
| E96/0 | Yes | 1019993.4 | Yes | No | 0.0 | No | 0.0 | Yes | 6675.6 | Late gametocytes only |
| E96/07 | Yes | 101498.0 | Yes | No | 0.0 | Yes | 9830.5 | Yes | 56989.1 | Early and late gametocytes |
| E96/21 | Yes | 107921.7 | No | No | 0.0 | No | 0.0 | No | 0.0 | pfs16 negative |
| E98/0 | Yes | 14957978.5 | No | No | 0.0 | Yes | 385.2 | No | 0.0 | pfs16 negative |
| E98/07 | Yes | 1162.4 | No | No | 0.0 | No | 0.0 | No | 0.0 | pfs16 negative |
| E98/14 | Yes | 1361.7 | No | No | 0.0 | No | 0.0 | No | 0.0 | pfs16 negative |
| E98/21 | Yes | 734.6 | No | No | 0.0 | No | 0.0 | No | 0.0 | pfs16 negative |
| E100/0 | Yes | 4897176.3 | Yes | Yes | 161.4 | Yes | 3341.1 | Yes | 33.2 | Early and late gametocytes |
| E100/07 | Yes | 1071652.1 | Yes | Yes | 10553.1 | Yes | 30994.7 | Yes | 24519.8 | Early and late gametocytes |
| E100/14 | Yes | 404063.6 | Yes | Yes | 10158.0 | Yes | 1138.8 | Yes | 363969.9 | Early and late gametocytes |
| E102/0 | Yes | 35490718.8 | Yes | Yes | 23.7 | Yes | 168.0 | No | 0.0 | Early gametocytes only |
| E102/14 | Yes | 105662.5 | Yes | Yes | 7.8 | No | 0.0 | Yes | 36519.3 | Early and late gametocytes |
| E102/21 | Yes | 739552.1 | Yes | Yes | 3.3 | No | 0.0 | Yes | 4137.4 | Early and late gametocytes |
| E103/0 | Yes | 101420968.8 | Yes | Yes | 3835.6 | Yes | 8141.9 | Yes | 2314.3 | Early and late gametocytes |
| E103/07 | Yes | 181.0 | No | No | 0.0 | Yes | 2982.6 | No | 0.0 | pfs16 negative |
| E103/14 | Yes | 79.3 | Yes | Yes | 49.1 | Yes | 711.2 | No | 0.0 | Early gametocytes only |
| E103/21 | Yes | 165.4 | No | Yes | 405.6 | No | 0.0 | Yes | 1762.5 | pfs16 negative |
| E104/0 | Yes | 20047294.9 | Yes | Yes | 199.5 | Yes | 3316.6 | No | 0.0 | Early gametocytes only |
| E104/07 | Yes | 671.5 | No | No | 0.0 | No | 0.0 | No | 0.0 | pfs16 negative |
| E104/14 | Yes | 138872.3 | Yes | Yes | 69.0 | No | 0.0 | Yes | 58189.8 | Early and late gametocytes |
| E104/21 | Yes | 2009.7 | Yes | Yes | 28.7 | No | 0.0 | Yes | 11652.5 | Early and late gametocytes |
| E105/0 | Yes | 943950.1 | Yes | No | 0.0 | No | 0.0 | Yes | 1077.6 | Late gametocytes only |
| E105/07 | Yes | 2291.0 | Yes | No | 0.0 | Yes | 16718.3 | No | 0.0 | Early gametocytes only |
| E105/14 | Yes | 2910.9 | Yes | No | 0.0 | Yes | 50075.5 | Yes | 62106.1 | Early and late gametocytes |
| E105/21 | Yes | 2191.9 | Yes | No | 0.0 | Yes | 1116.7 | Yes | 110932.4 | Early and late gametocytes |
| E106/0 | Yes | 9702050.8 | Yes | Yes | 196.3 | No | 0.0 | Yes | 1218.5 | Early and late gametocytes |
| E106/07 | Yes | 349.8 | Yes | Yes | 3.3 | No | 0.0 | No | 0.0 | Early gametocytes only |
| E106/14 | Yes | 76.5 | No | No | 0.0 | No | 0.0 | No | 0.0 | pfs16 negative |
| E106/21 | Yes | 59.5 | No | No | 0.0 | No | 0.0 | No | 0.0 | pfs16 negative |

PID Patient ID

| | |
|---|---|
| Total parasite | Presence of any parasite stage |
| Q.Total parasite | Total parasite quantity/mL of blood |
| Any Gametocyte | Presence of any gametocyte stage (pfs16) |
| Early (*pfpeg4*) | Presence of early gametocyte (*pfpeg4*) |
| Q.Early (*pfpeg4*) | Early gametocyte quantity (*pfpeg4*)/mL of blood |
| Early (*pfg27*) | Presence of early gametocyte (*pfg27*) |
| Q. Early (*pfg27*) | Early gametocyte quantity (*pfg27*)/mL of blood |
| Late (*pfs25*) | Presence of late gametocyte (*pfs25*) |
| Q.Late (*pfs25*) | Late gametocyte quantity (*pfs25*)/mL of blood |

Coloured cells represents samples with detectable early gametocytes markers expression, though their quantities are below the limit of quantification (the lowest ivcDNA concentration that detectable in all of the experiments and falls within the log-linear relationship of the RT-qPCR assay)

**Table S5: Raw data for volunteers samples**

| ID | Day | Tparasite.mlb | MGCTmlbl | EGCTpeg4.mlbl | EGCT27.mlbl | $C_T$40S |
|---|---|---|---|---|---|---|
| S035 | 7.0 | 15721 | 0.0 | 0.0 | 0.0 | 34.9 |
| S035 | 7.5 | 4599 | 270.1 | 0.0 | 0.0 | 35.3 |
| S035 | 8.0 | 42190 | 0.0 | 0.0 | 0.0 | 37.3 |
| S035 | 8.1 | 91857 | 0.0 | 0.0 | 139.2 | 35.8 |
| S035 | 8.2 | 126694 | 167.1 | 105.9 | 247.2 | 34.0 |
| S035 | 8.5 | 131359 | 175.3 | 850.3 | 0.0 | 32.2 |
| S035 | 8.7 | 157358 | 164.4 | 144.3 | 0.0 | 32.4 |
| S035 | 8.8 | 45510 | 0.0 | 218.4 | 112.5 | 35.2 |
| S035 | 9.0 | 7087 | 0.0 | 0.0 | 0.0 | 37.1 |
| S035 | 9.3 | 1281 | 0.0 | 0.0 | 0.0 | 39.4 |
| S042 | 7.0 | 14396 | 0.0 | 196.2 | 0.0 | 37.9 |
| S042 | 8.0 | 109920 | 0.0 | 319.0 | 0.0 | 35.7 |
| S042 | 8.1 | 289620 | 170.7 | 461.6 | 302.1 | 34.5 |
| S042 | 8.2 | 333068 | 279.4 | 363.0 | 157.6 | 32.9 |
| S042 | 8.5 | 227850 | 247.1 | 271.8 | 370.6 | 34.3 |
| S042 | 8.7 | 114926 | 80.0 | 618.7 | 285.9 | 35.5 |
| S042 | 8.8 | 68042 | 190.7 | 203.0 | 138.3 | 33.1 |
| S042 | 9.0 | 6634 | 0.0 | 486.6 | 147.1 | 38.8 |
| S042 | 9.3 | 836 | 0.0 | 0.0 | 0.0 | 41.3 |

**ID** Volunteer ID
**Day** Day of follow up
**Tparasite.mlb** Total parasite density/ mL of blood
**MGCTmlbl** Late gametocytes/ mL of blood
**EGCTpeg4.mlbl** Early gametocytes (pfpeg4)/mL of blood
**EGCT27.mlbl** Early gametocytes (pfg27)/mL of blood
**$C_T$40S** PCR cycle threshold of pf40S (endogenous control gene)

40S RT-qPCR was used to detect any DNA carryover in the purified RNA samples (reverse transcriptase-free sample), cycle threshold of pf40S was undetermined in all of the samples above indicating no DNA contamination.

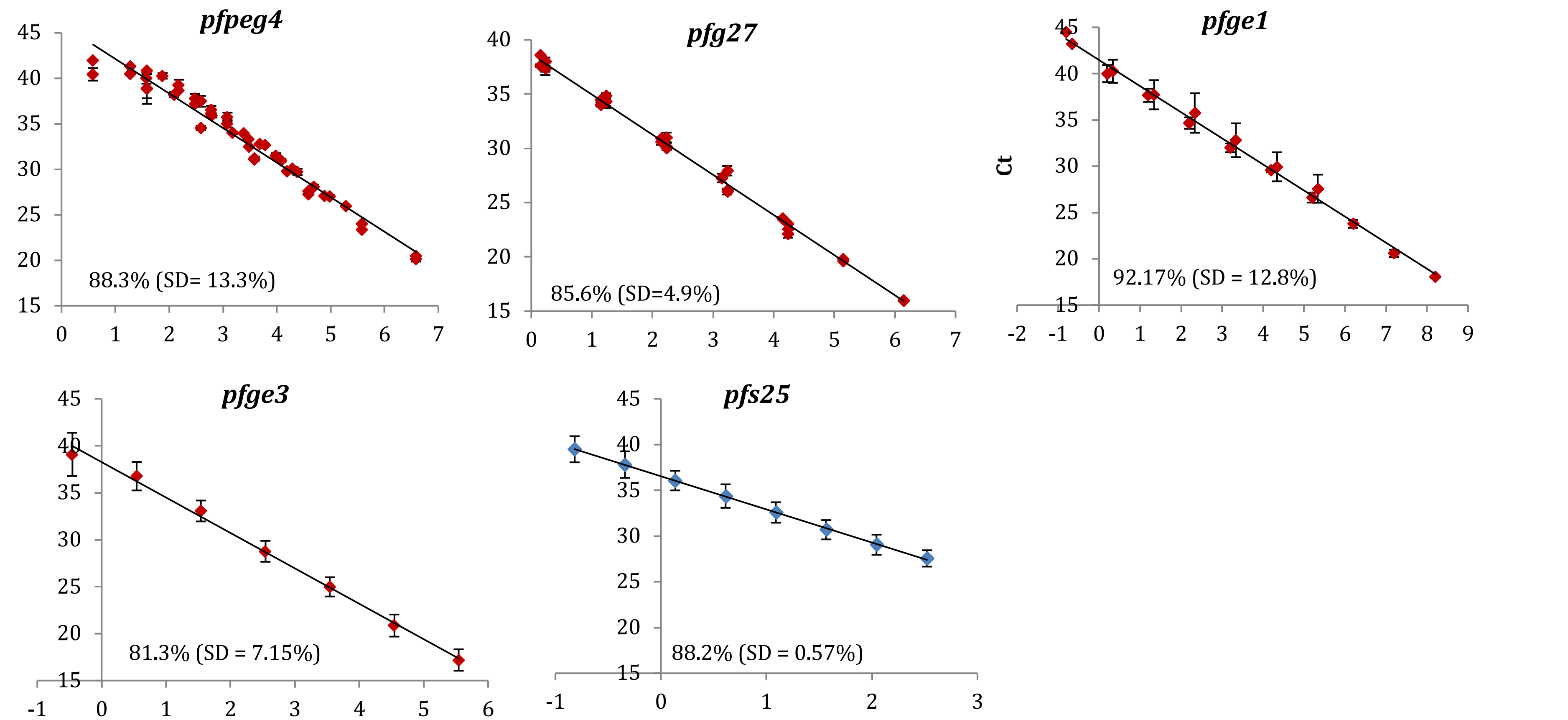


**Figure S1: *pfpeg4*, *pfg27*, *pfge1*, *pfge3* and *pfs25* quantification standard curves.** Standard curve generated from serially diluted ivcDNA (methods section 2.6.4). The average efficiency of the standard curves and standard deviation (SD) are given within each plot. Y axis represents the cycle threshold ($C_T$) at which the change in the probe fluorescence was first detectable. X axis represents log10 of the ivcDNA concentration (transcript copies/µL of ivcDNA) for *pfpeg4*, *pfg27*, *pfge1* and *pfge3* standard curves. For *pfs25* the x axis represents mature gametocytes/µL of cDNA. Error bars represent the standard deviation of the $C_T$ replicates.

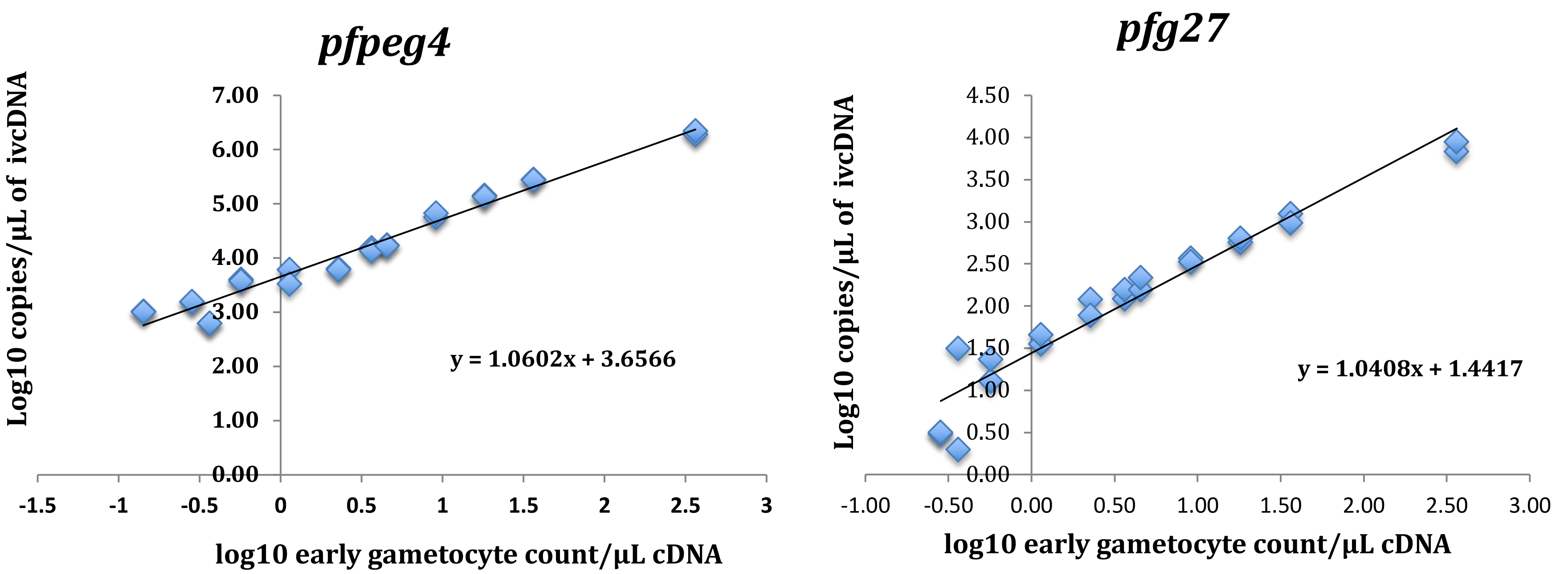


**Figure S2: Conversion from transcript number to gametocytes count using *pfpeg4* and *pfg27* genes.** Standard curve generated from serially diluted ivcDNA (Y axis) and purified stage II gametocytes (X axis). No conversion curves were produced for the other early gametocytes markers because proved less specificity (*pfge1* and *pfgexp5*) or lower abundance (*pfge3*).

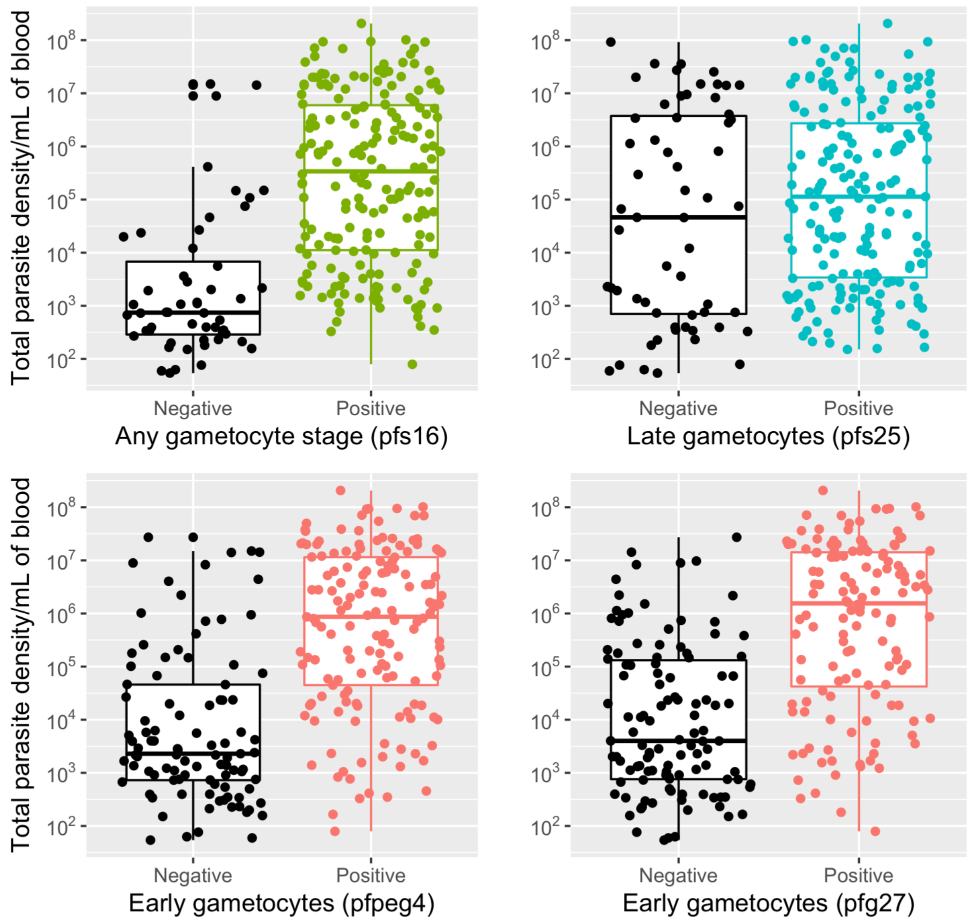


**Figure S3:** Detection of gametocytes stages across a wide range of parasitaemia.
